# Efficacy of Repeated-Dose Albendazole/ Mebendazole Regimens for Soil-Transmitted Helminth Infections in Paediatric Population: A Systematic Review and Meta-Analysis

**DOI:** 10.64898/2026.10.09.26365109

**Authors:** Nishant Singh, Priyanka Sahu, Vaibhav Singh, Prem Prakash Bharti, Arshiya Masood siddiqui, Arun Kumar

**Author notes:** CORRESPONDING AUTHOR: Dr. Nishant Singh, MD, Assistant Professor, Department of Community Medicine, Government Medical College, Badaun, Uttar Pradesh, India. These authors contributed equally to this work.

## Abstract

**Background:** The primary approach to controlling soil-transmitted helminths (STH) is preventive chemotherapy using single-dose benzimidazoles. However, single-dose regimens frequently result in suboptimal clearance against species like Trichuris trichiura and hookworm. This systematic review and meta-analysis evaluated the comparative efficacy of repeated-dose versus standard single-dose albendazole or mebendazole regimens in paediatric populations.

**Methods:** Registered prospectively on PROSPERO (CRD420261491616), a comprehensive search of PubMed, Embase, Cochrane Library, and Google Scholar was conducted from database inception up to September 1, 2026, restricted to English-language publications. The search identified clinical trials comparing repeated-dose against single-dose albendazole or mebendazole in paediatric STH infections. Primary outcomes were cure rate (CR) and egg reduction rate (ERR). Random-effects models synthesized pooled odds ratios (OR) and mean differences (MD). Risk of bias was assessed using the Cochrane RoB 2 tool.

**Results:** Five randomized clinical trials (N = 1,718 participants) met the inclusion criteria. Overall, repeated dosing demonstrated significant superiority in cure rate (pooled OR = 4.47, 95% CI: 1.58–12.67; I² = 92%) and egg reduction rate (pooled MD = 0.17, 95% CI: 0.01–0.32; I² = 87%).

**Discussion:** Repeated-dose benzimidazole regimens offer superior overall therapeutic efficacy compared to standard single-dose administration in children. However, this therapeutic advantage is drug-specific: multi-dose albendazole consistently enhances parasite clearance across trial settings, whereas multi-dose mebendazole exhibits variable performance and lacks conclusive superiority over single dosing.

**Conclusion:** Transitioning to targeted, multi-dose albendazole protocols—rather than mebendazole—may serve as an effective public health intervention to eliminate residual parasite reservoirs in high-transmission endemic regions.

**Author summary:** While current WHO guidelines recommend single-dose preventive chemotherapy for soil-transmitted helminths, standard regimens often fail to achieve optimal clearance for Trichuris trichiura and hookworms. Our meta-analysis of randomized clinical trials (N = 1,718 children) demonstrates that repeated-dose regimens significantly improve cure rates (pooled OR = 4.47; post-sensitivity OR = 2.60) and egg reduction rates (pooled MD = 0.17; post-sensitivity MD = 0.08), driven specifically by albendazole (cure rate OR = 3.10, I-squared = 7%; egg reduction rate pooled MD = 0.07, 95% CI: 0.03–0.11, I-squared = 0%, Z = 3.29, P = 0.001) rather than mebendazole. This work provides key empirical evidence that shifting public health policy from single-dose mass drug administration to targeted, multi-dose albendazole in high-transmission hotspots can effectively eliminate persistent parasite reservoirs and prevent therapeutic failure.

## Introduction

Soil-transmitted helminths (STH) harm over two billion people globally ^1, 2^. They are primarily found in preschoolers and school-age children and are extensively endemic in the tropics^3, 4^ as well as in pregnant women^5^. Infections are associated with poor sanitation and hygiene and classed as neglected tropical illnesses.^6^ These infections cause severe morbidity, including anemia^7, 8^, stunting^9^, and iron and vitamin A deficiencies**^10–13^**. Furthermore, helminth infections can lower the host immune response to environmental and parasite antigens, as well as to vaccinations^14–16^. The foundation of the STH control strategy has been preventive chemotherapy. However, successful management requires increased access to potable water, proper sanitation, vector control, and health education to expedite progress toward eradication^17–19^. To tackle soil-transmitted helminths (STHs), the World Health Organization advises administering benzimidazole treatments once or twice a year to toddlers (ages 1 to 4) and school-aged children (ages 5 to 14) residing in regions where infection rates top 20%. The standard preventative dosage includes either albendazole (200 mg for toddlers aged 1–2, or 400 mg for everyone older) or a single 500 mg dose of mebendazole^20^. Mass drug administration (MDA) campaigns favor using albendazole (ALB) or mebendazole (MBZ) because they are administered as a single-dose tablet, making patient compliance much easier to achieve^21^. Since 2003, Uganda has been running a mass drug administration (MDA) campaign using albendazole and mebendazole, deworming millions of children every year^22^. A single 400 mg dose of albendazole is rarely enough to tackle Trichuris trichiura. In fact, trial data from Bangladesh showed that clearing 80% of egg counts required stretching treatment across three consecutive days.^23^.

## Rationale

This systematic review and meta-analysis was conducted to quantify the exact comparative therapeutic advantage—measured by cure rates and egg reduction rates—of repeated-dose versus single-dose albendazole/ mebendazole regimens in pediatric population, providing pooled evidence to inform and optimize deworming policies.

## Method and Search strategy

The review protocol was registered prospectively on the International Prospective Register of Systematic Reviews (PROSPERO) under registration number CRD420261491616. We conducted a systematic literature search across PubMed, Embase, the Cochrane Library, and Google Scholar from database inception up to September 1, 2026. The search was restricted to studies published in the English language only, with no restrictions on publication date. To identify relevant studies, we built a comprehensive search string combining Medical Subject Headings (MeSH) and free-text terms using Boolean operators: (“child” OR “preschool” OR “pediatric” OR “infant”) AND (“soil transmitted helminth” OR “STH” OR “Ascaris” OR “Trichuris” OR “hookworm”) AND (“albendazole” OR “mebendazole” OR “benzimidazole”) AND (“single dose” OR “single-dose” OR “multi dose” OR “repeated dose”). The database searches yielded a total of 1,448 records. We uploaded 1,448 identified records into Rayyan, removed 36 duplicates, and screened the titles and abstracts of the remaining 1,412 unique studies. Two independent reviewers evaluated all records against predefined criteria, leading to the exclusion of 1,386 studies. The reasons for exclusion at this initial stage were wrong outcomes (n = 527), wrong intervention or drug (n = 431), wrong population (n = 279), wrong publication (n = 93), and wrong study design (n = 56). We then retrieved and reviewed the full texts of the remaining 26 studies. Of these, 21 were excluded after detailed evaluation due to wrong outcomes (n = 10), wrong population (n = 8), or wrong study design (n = 3). Ultimately, 5 studies met all inclusion criteria and were included in the final data extraction and quantitative meta-analysis. **(Figure 1)**

**Figure 1.**
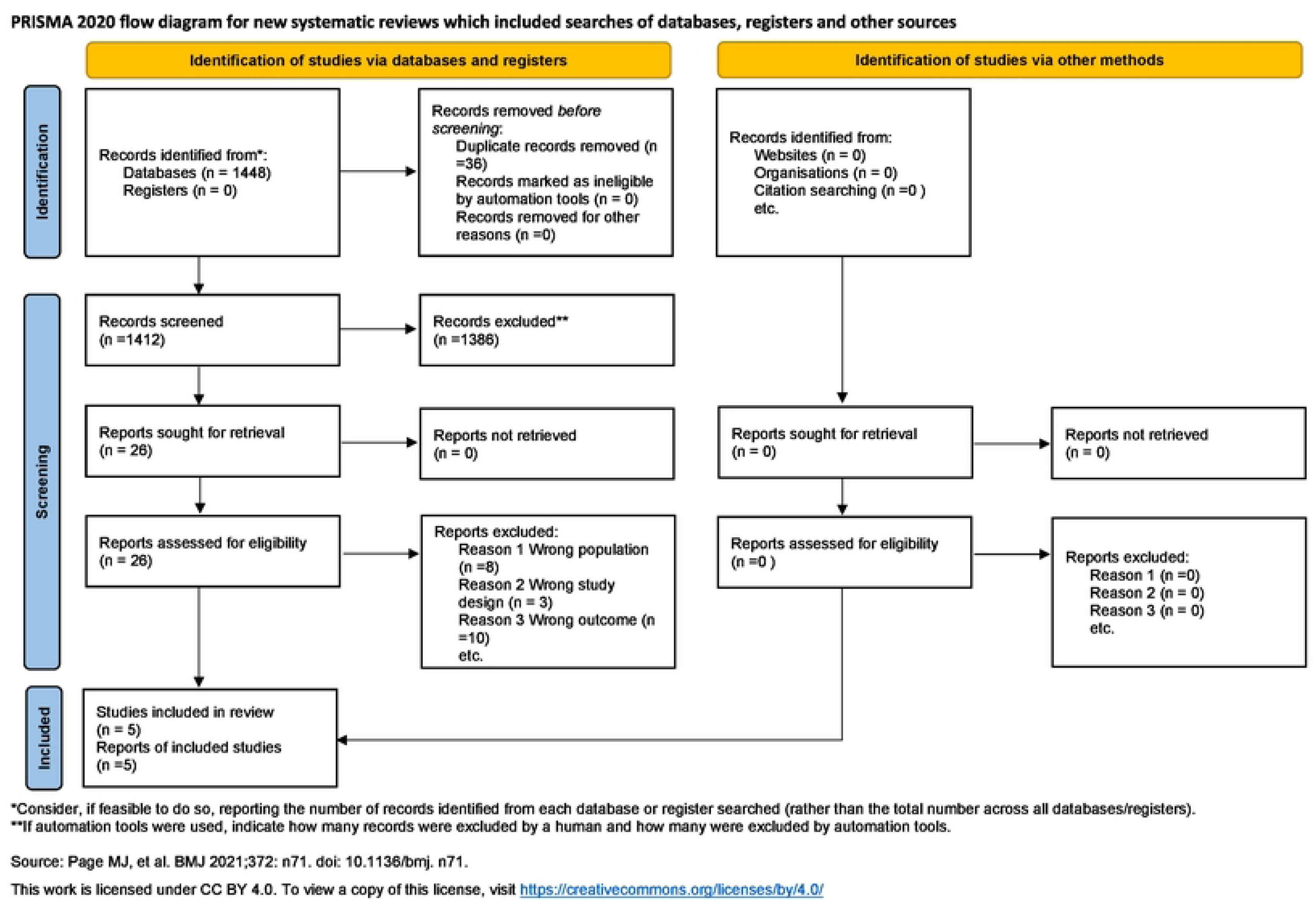
PRISMA flow diagram showing the screening and selection process

## Eligibility Criteria

Only studies that strictly adhere to the predefined PICO framework were included in this review. This systematic review strictly includes clinical trials evaluating pediatric populations with parasitologically confirmed soil-transmitted helminth infections (Ascaris lumbricoides, Trichuris trichiura, or hookworms). Eligible trials must compare repeated-dose regimens (multi-day or consecutive daily schedules) of albendazole or mebendazole directly against single-dose regimens of the same agent, reporting cure rate (CR) or egg reduction rate (ERR). Studies focusing exclusively on adults, utilizing co-interventions with other anthelmintics, lacking a concurrent single-dose control group, or employing observational, mathematical, or secondary review designs are excluded.

## Data abstraction and analysis

Data extraction was conducted independently by two reviewers using a standardized electronic template, with discrepancies resolved through consensus or third-party arbitration. The template systematically captured study metadata (first author, publication year, title, journal, DOI/PMID, country, language, funding source, conflicts of interest, trial registration number, publication status) **(Table 1)**, study design characteristics (design type, parallel group structure, number of arms, unit of randomization, follow-up duration, inclusion/exclusion criteria, setting), and baseline demographics (sample sizes randomized and analyzed, mean age, age range, sex distribution, baseline disease severity, diagnostic criteria, comorbidities, socioeconomic factors). Detailed intervention and comparator protocols were recorded, including drug names, intervention type, dose intensity, dosing frequency, treatment duration, route of administration, provider type, co-interventions, and adherence measurements. Primary efficacy outcome parameters—specifically Cure Rate (CR) and Egg Reduction Rate (ERR)—were extracted as dichotomous event data (events and sample sizes) and continuous data (means, standard deviations, sample sizes) across treatment and control arms.

**Table 1.** Identification detail of included studies.

| Study ID | First Author | Year of Publication | Title | Journal | DOI / PMID | Country / Countries | Funding Source | Conflict of Interest Declared | Trial Registration Number |
| --- | --- | --- | --- | --- | --- | --- | --- | --- | --- |
| **Adeg nika et al. (2014)**<sup>24</sup> | Ayola A. Adeg nika | 2014 | Randomized, Controlled, Assessor-Blind Clinical Trial To Assess the Efficacy of Single-versus Repeat ed-Dose Albendazole To Treat *Ascaris lumbricoides*, *Trichuris trichiura*, and Hookworm Infection | Antimicrobial Agents and Chemotherapy | 10.1128/AAC.01317-13 | Gabon | EDCTP, Deutsche Forschungsgemeinschaft (DFG), European Union (IDEA and TheSchistoVac projects) | None declared | NCT01192802 |
| **Kim et al. (2023)**<sup>25</sup> | Eun Seok Kim | 2023 | The impact of dual-versus single-dosing and fatty food co-administration on albendazole efficacy against hookworm among children n in Mayuge district, Uganda : Results from a 2x2 factorial randomised controlled trial | PLOS Neglected Tropical Diseases | 10.1371/journal.pntd.0011439 | Uganda | Korea International Cooperation Agency (KOICA), World Vision | No | PACTR202202738940158 |
| **Namwanje et al. (2011)**<sup>26</sup> | Harriet Namwanje | 2011 | Efficacy of single and double doses of albendazole and mebendazole alone and in combination in the treatment of *Trichuris trichiura* in school-age children in Uganda | Transactions of the Royal Society of Tropical Medicine and Hygiene | 10.1016/j.trstmh.2011.07.009 | Uganda | DBL-Centre for Health Research and Development, University of Copenhagen | None declared | NCT01050452 |
| **Steinmann et al. (2011)**<sup>27</sup> | Peter Steinmann | 2011 | Efficacy of Single-Dose and Triple-Dose Albendazole and Mebendazole against Soil-Transmitted Helminths and *Taenia* spp.: A Randomized Controlled Trial | PLoS ONE | 10.1371/journal.pone.0025003 | People's Republic of China | Stanley Thomas Johnson Foundation, Swiss National Science Foundation, Novartis Foundation, Ministry of Science and Technology (China), UNICEF/UNDP/World Bank/WHO TDR | PS supported by Novartis Foundation | ISRCTN47375023 |
| **Eshetu et al. (2020)**<sup>28</sup> | Tegegne Eshetu | 2020 | Efficacy of a single dose versus a multiple dose regimen of Mebendazole against hookworm infections among school children: a randomized open-label trial | BMC Infectious Diseases | 10.1186/s12879-020-05097-1 | Ethiopia | University of Gondar | No | PACTR201911466695052 |

## Risk of Bias Assessment of Included Studies

The methodological quality of five randomized controlled trials was evaluated across five domains using the Cochrane Risk of Bias 2 (RoB 2) tool. Overall, two studies (Adegnika et al., 2014^24^; Kim et al., 2023^25^) demonstrated a low risk of bias across all domains (D1–D5). Conversely, three studies (Namwanje et al., 2011^26^; Steinmann et al., 2011^27^; Eshetu et al., 2020^28^) were categorized as having “some concerns” overall. Across specific domains, selection of reported results (D5) was low risk in all five studies. Measurement of the outcome (D4) was low risk in four studies, with Namwanje et al. (2011)^26^ presenting insufficient details (“no information”). Missing outcome data (D3) was low risk in three studies, while Steinmann et al. (2011)^27^ and Namwanje et al. (2011)^26^ exhibited “some concerns.” Both the randomization process (D1) and deviations from intended interventions (D2) were judged low risk in Adegnika et al. (2014)^24^, Kim et al. (2023)^25^, and Steinmann et al. (2011)^27^, but raised “some concerns” in Namwanje et al. (2011)^26^ and Eshetu et al. (2020)^28^. **(Figure 2)**

**Figure 2.**
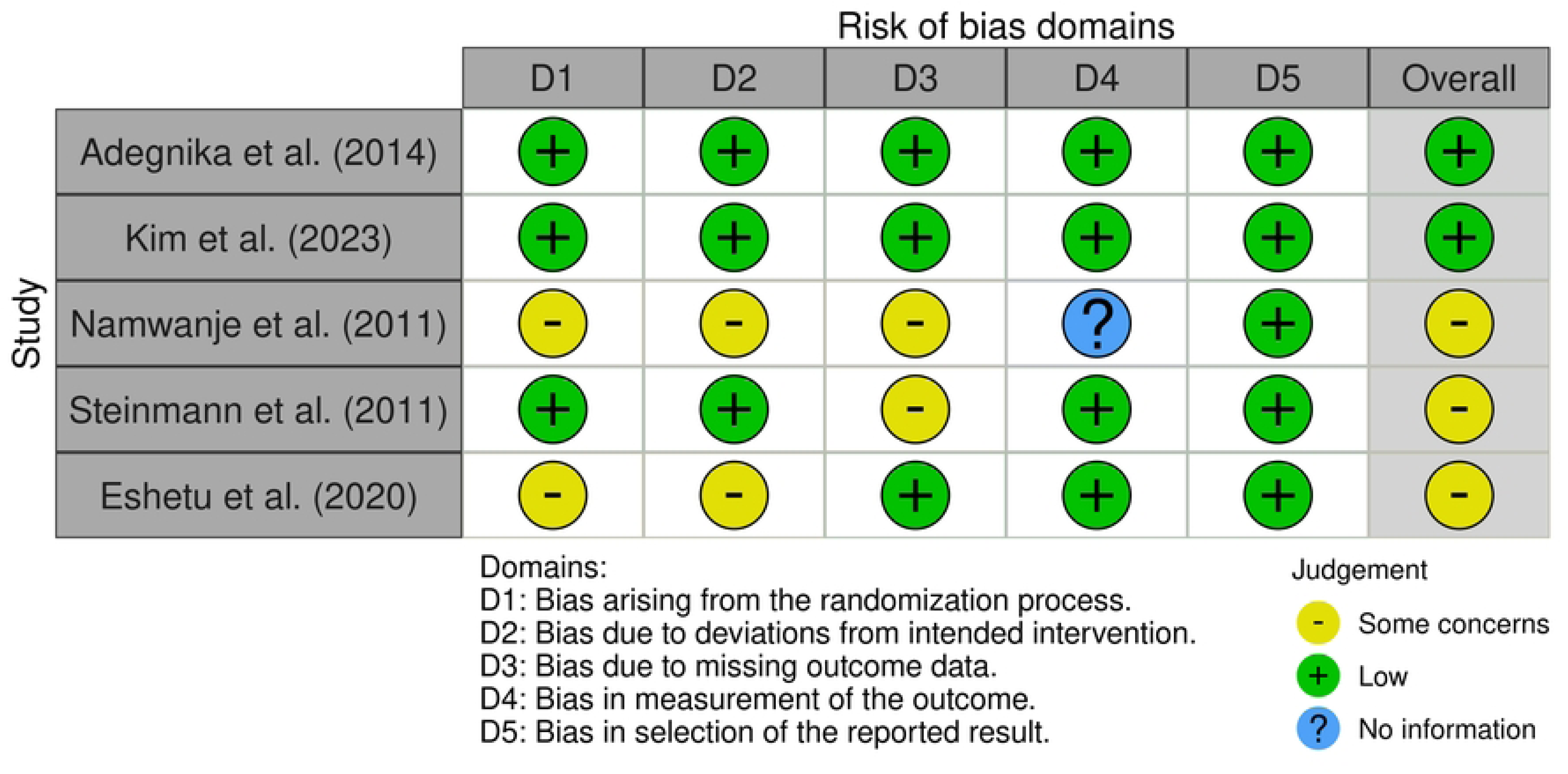
Risk of Bias Assessment of Included Studies

## Result

Repeated-dose albendazole/mebendazole demonstrated statistically significant superiority over single-dose therapy for cure rates, yielding a pooled odds ratio of 4.47 (95% CI: 1.58 to 12.67; Z = 2.82, P = 0.005) under a random-effects model using Restricted Maximum-Likelihood (REML) tau-squared estimation and Wald-type confidence intervals. The meta-analysis synthesized five studies with 889 experimental participants (619 cure events) and 829 control participants (429 cure events). Individual trial point estimates consistently favored repeated dosing: Eshetu et al. (2020)^28^ reported an OR of 55.13 (95% CI: 11.92 to 255.00; weight 15.5%), Kim et al. (2023)^25^ an OR of 5.07 (95% CI: 1.66 to 15.53), Adegnika et al. (2014)^24^ an OR of 3.71 (95% CI: 2.13 to 6.46), Steinmann et al. (2011)^27^ an OR of 2.20 (95% CI: 1.59 to 3.05; weight 22.9%), and Namwanje et al. (2011)^26^ an OR of 1.67 (95% CI: 0.85 to 3.28). Substantial statistical heterogeneity was observed across study populations, indicated by an I-squared value of 92%, a Chi-squared statistic of 20.81 (df = 4, P = 0.0003), and a Tau-squared variance of 1.21. **(Figure 3)**

**Figure 3.**
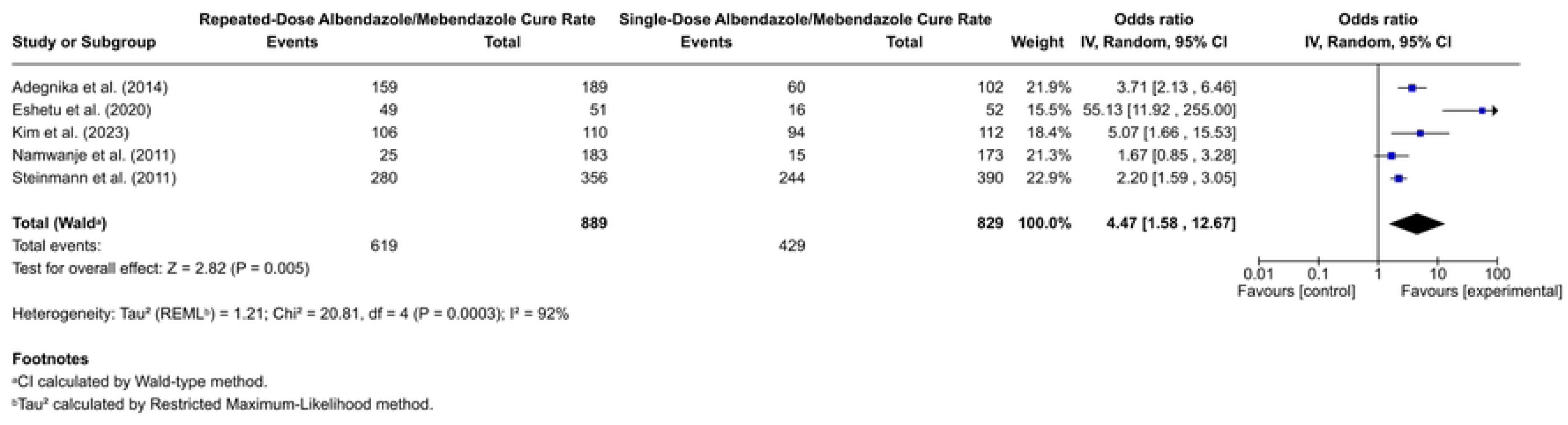
Forest Plot Comparing Cure Rate of Repeated-Dose versus Single-Dose Albendazole/Mebendazole Treatment

This sensitivity analysis re-evaluates repeated-dose versus single-dose albendazole/mebendazole treatment after excluding the outlier by Eshetu et al. (2020) (0.0% weight). Across the four remaining studies (838 repeated-dose participants with 570 cures; 777 single-dose participants with 413 cures), study weights were reallocated to Steinmann et al. (2011)^27^ at 42.0%, Adegnika et al. (2014)^24^ at 26.9%, Namwanje et al. (2011)^26^ at 21.1%, and Kim et al. (2023)^25^ at 10.0%. Individual odds ratios favored repeated doses, ranging from 1.67 (95% CI: 0.85–3.28) in Namwanje et al.^26^ to 5.07 (95% CI: 1.66–15.53) in Kim et al.^25^, alongside 2.20 (95% CI: 1.59–3.05) in Steinmann et al.^27^ and 3.71 (95% CI: 2.13– 6.46) in Adegnika et al.^24^ The pooled odds ratio confirmed significant superiority for repeated-dose therapy at 2.60 (95% CI: 1.76–3.83; Z = 4.83, P < 0.00001). Excluding the outlier reduced I-squared heterogeneity from 92% to 43% (Chi-squared = 5.42, df = 3, P = 0.14; Tau-squared = 0.07), confirming robust therapeutic advantage of repeated-dose. **(Figure 4)**

**Figure 4.**
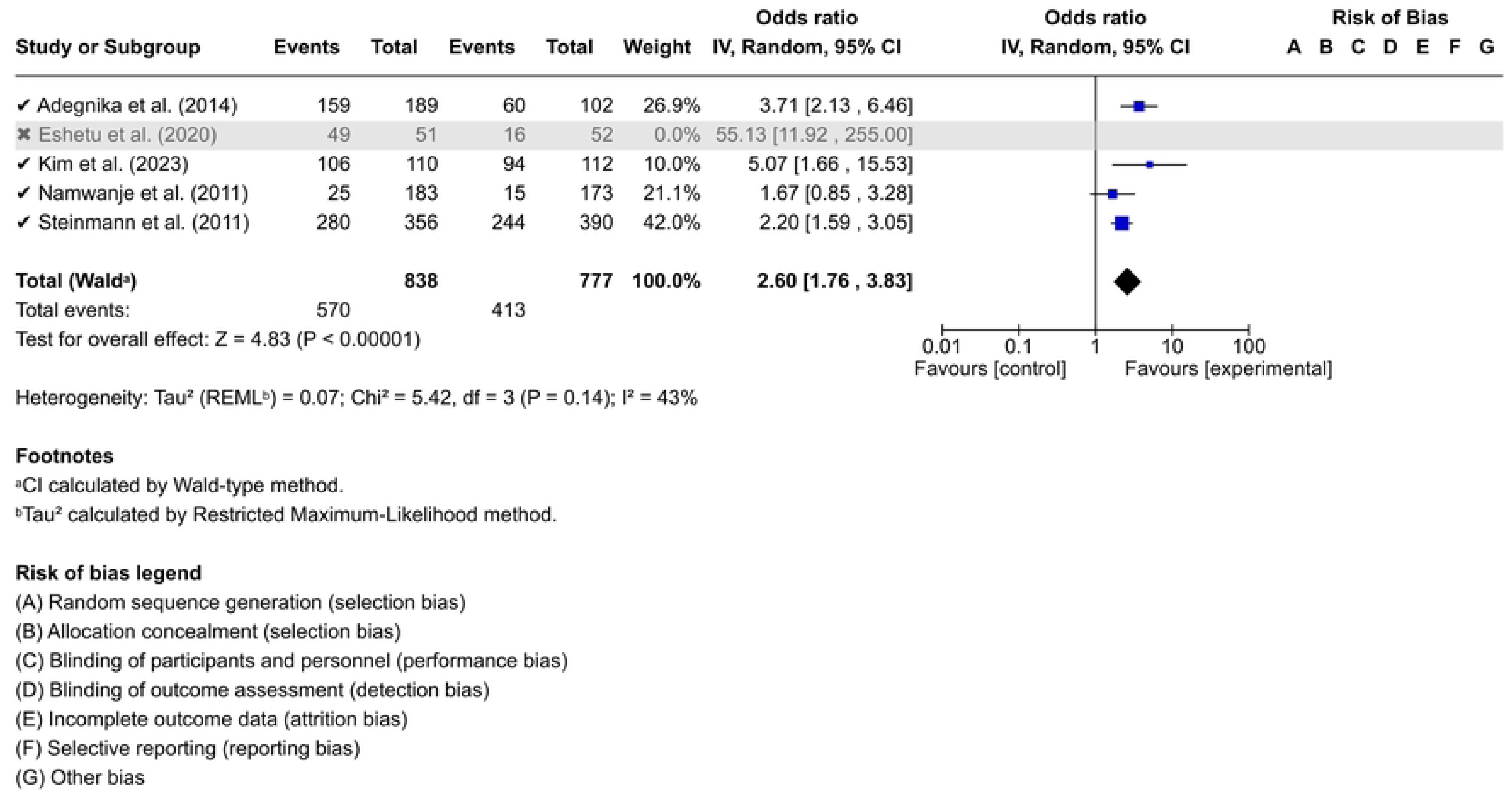
Forest plot comparing cure rates of repeated-dose versus single-dose albendazole/mebendazole after sensitivity analysis

Repeated-dose Albendazole or Mebendazole demonstrates significantly higher egg reduction rates than single-dose treatment across four studies (706 repeated-dose vs. 656 single-dose participants). The pooled mean difference is 0.17 (95% CI: 0.01, 0.32; Z = 2.13, P = 0.03). Individual study mean differences and random-effects model weights are: Adegnika et al. at^24^ 0.39 (95% CI: 0.05, 0.73; 12.8% weight), Eshetu et al.^28^ at 0.31 (95% CI: 0.18, 0.44; 27.1% weight), Kim et al.^25^ at 0.03 (95% CI: -0.09, 0.15; 27.6% weight), and Steinmann et al.^27^ at 0.08 (95% CI: 0.05, 0.11; 32.5% weight). Significant heterogeneity was present across trials (Chi² = 16.32, df = 3, P = 0.0010; I² = 87%; Tau² = 0.02). **(Figure 5)**

**Figure 5.**
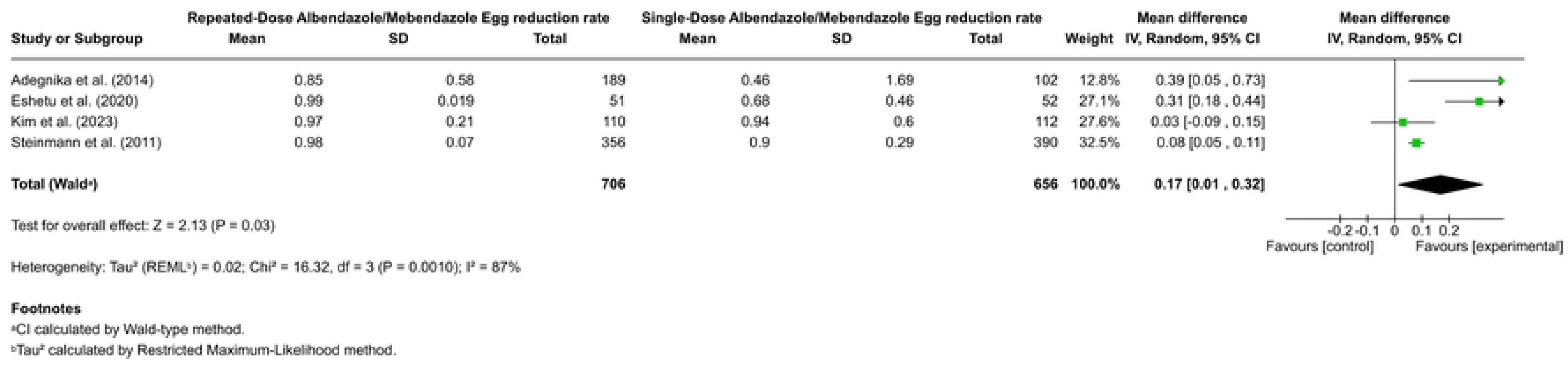
Forest plot comparing egg reduction rates of repeated-dose versus single-dose albendazole/mebendazole

Following the sensitivity analysis excluding Eshetu et al. (2020)^28^, three studies (Adegnika et al. 2014^24^ [0.7% weight], Kim et al. 2023^25^ [5.9% weight], and Steinmann et al. 2011^27^ [93.4% weight]) were synthesized, comprising 1,259 participants (655 experimental repeated-dose, 604 control single-dose). Using a random-effects inverse variance model with Restricted Maximum-Likelihood estimation, the pooled mean difference in egg reduction rate was 0.08 (95% CI: 0.05 to 0.11), significantly favoring the repeated-dose regimen over single-dose treatment (Z = 5.42, P < 0.00001). The retained studies demonstrated complete homogeneity (Chi² = 3.92, df = 2, P = 0.14, Tau² = 0.00, I² = 0%), confirming that repeated dosing with albendazole or mebendazole yields a statistically significant and highly consistent improvement in egg reduction rate. **(Figure 6)**

**Figure 6.**
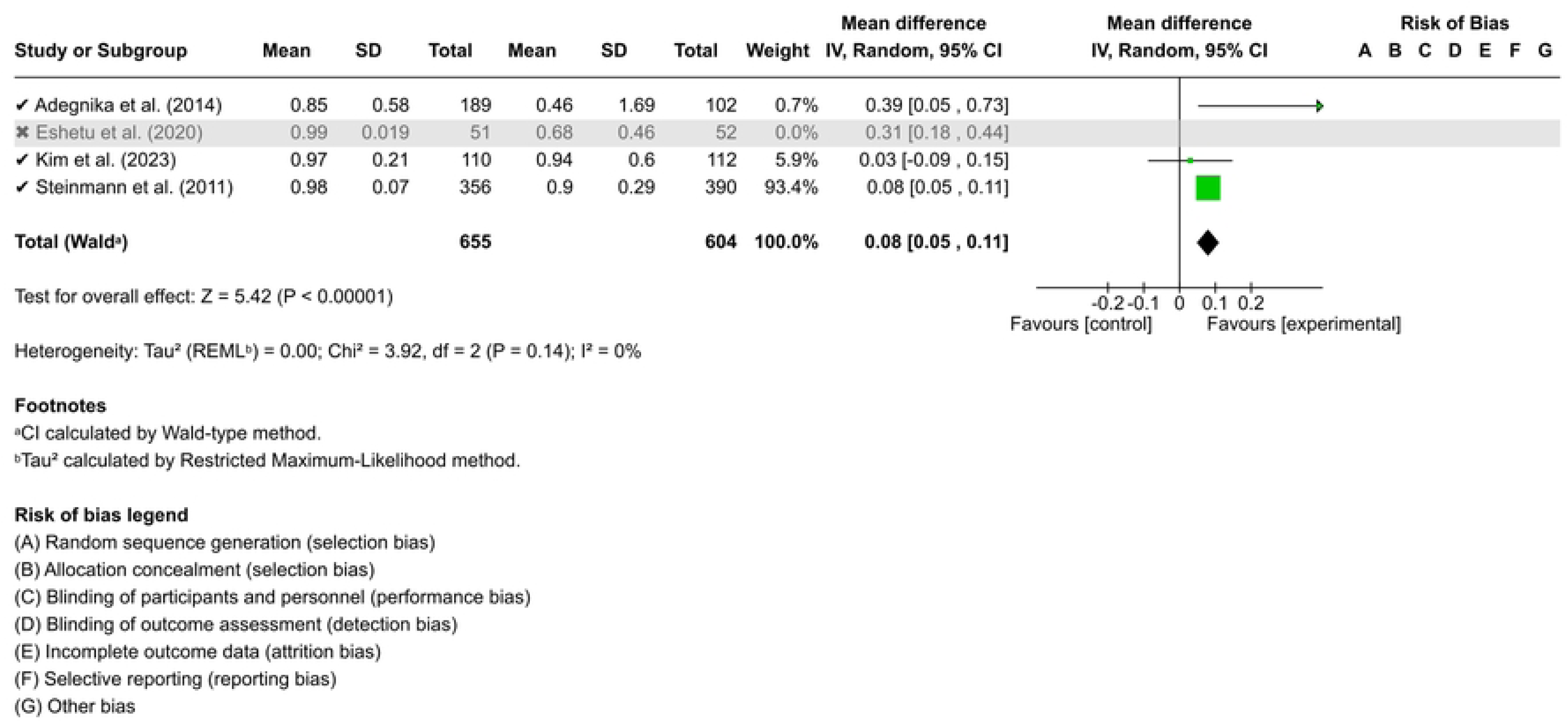
Forest plot comparing egg reduction rates of repeated-dose versus single-dose albendazole/mebendazole after sensitivity analysis

Repeated-dose albendazole demonstrated superior cure rate efficacy compared to single-dose albendazole across four studies: Adegnika et al. (2014)^24^ (OR = 3.71, 95% CI: 2.13–6.46; weight = 36.8%), Kim et al. (2023)^25^ (OR = 5.07, 95% CI: 1.66–15.53; weight = 9.9%), Namwanje et al. (2011)^26^ (OR = 3.36, 95% CI: 1.15–9.82; weight = 10.8%), and Steinmann et al. (2011)^27^ (OR = 2.32, 95% CI: 1.39–3.86; weight = 42.5%). Across the trials, the repeated-dose group achieved 413 cure events among 539 participants, compared to 294 cure events among 495 participants in the single-dose group. A random-effects model yielded an overall pooled odds ratio of 3.10 (95% CI: 2.17–4.44), indicating more than three times higher odds of cure with repeated doses. This superiority was highly statistically significant (Z = 6.18, P < 0.00001). Heterogeneity was minimal across studies (Tau² = 0.01, Chi² = 2.42, df = 3, P = 0.49, I² = 7%), demonstrating strong consistency in treatment outcomes. **(Figure 7)**

**Figure 7.**
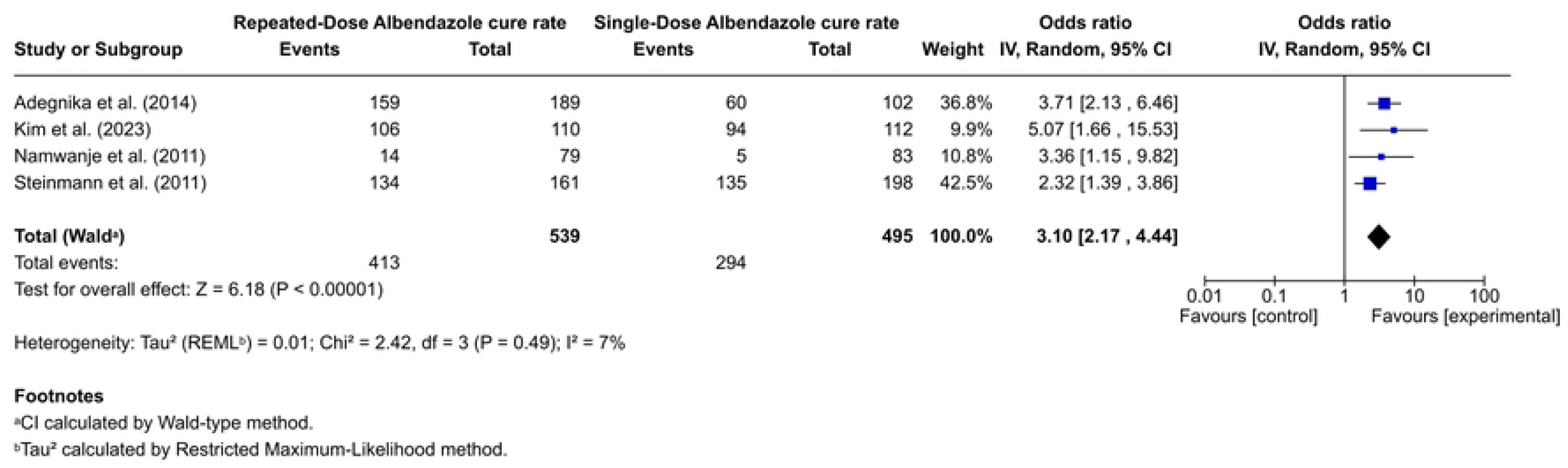
Forest plot comparing cure rates of repeated-dose versus single-dose albendazole

Across three studies involving 872 participants (460 repeated-dose, 412 single-dose), repeated-dose albendazole demonstrated superior efficacy over single-dose administration in achieving higher egg reduction rates. Individual study effect sizes consistently favored the repeated-dose regimen: Adegnika et al. (2014)^24^ observed a mean difference of 0.39 (95% CI: 0.05 to 0.73; weight = 1.5%), Kim et al. (2023)^25^ reported 0.03 (95% CI: -0.09 to 0.15; weight = 12.5%), and Steinmann et al. (2011)^27^ contributed the primary weight (86.0%) with a mean difference of 0.07 (95% CI: 0.03 to 0.11). Under a random-effects model, the overall pooled mean difference was 0.07 (95% CI: 0.03 to 0.11; Z = 3.29, P = 0.001), indicating a statistically significant benefit for repeated dosing. Statistical heterogeneity was negligible across the trials (Chi-squared = 3.88, df = 2, P = 0.14; Tau-squared = 0.00; I-squared = 0%). **(Figure 8)**

**Figure 8.**
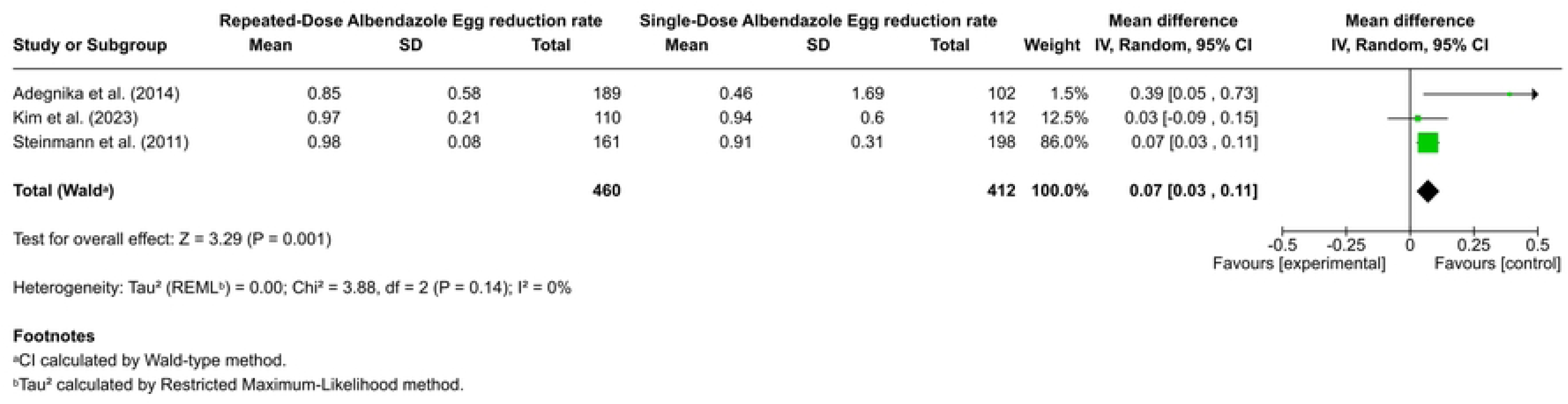
Forest plot comparing egg reduction rates of repeated-dose versus single-dose albendazole

This forest plot evaluates repeated-dose versus single-dose mebendazole for achieving cure across three studies totaling 684 participants (350 repeated-dose, 334 single-dose). Outcomes varied significantly across individual trials: Eshetu et al. (2020)^28^ demonstrated a substantial advantage for repeated dosing with an OR of 55.13 (95% CI: 11.92 to 255.00; weight: 30.9%; 49/51 vs. 16/52 events), Steinmann et al. (2011)^27^ showed a moderate, statistically significant benefit favoring repeated dosing with an OR of 2.27 (95% CI: 1.47 to 3.49; weight: 35.3%; 146/195 vs. 109/192 events), and Namwanje et al. (2011)^26^ indicated no significant difference with an OR of 0.95 (95% CI: 0.38 to 2.34; weight: 33.8%; 11/104 vs. 10/90 events). Overall, 206 cures occurred in the repeated-dose group compared to 135 in the single-dose group. Using a random-effects model (REML estimation, Wald-type CIs), the pooled odds ratio was 4.52 (95% CI: 0.44 to 46.24). This overall effect was not statistically significant (Z = 1.27, P = 0.20), and extreme heterogeneity was present across studies (Tau² = 3.94; Chi² = 20.19, df = 2, P < 0.0001; I² = 95%), rendering the pooled superiority of repeated dosing inconclusive. **(Figure 9)**

**Figure 9.**
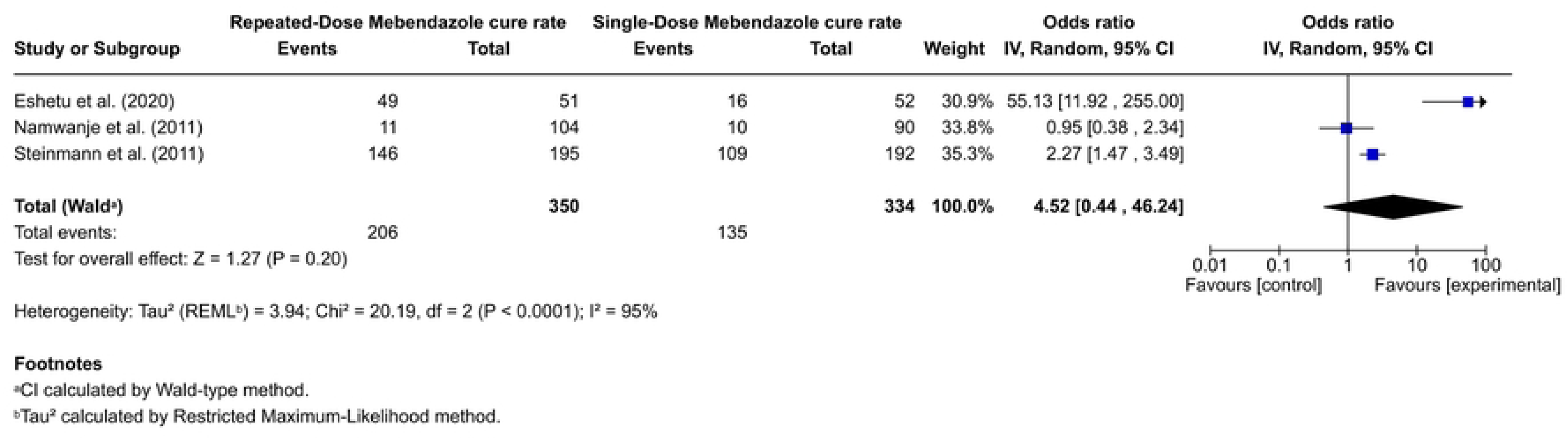
Forest plot comparing cure rates of repeated-dose versus single-dose mebendazole

This forest plot compares the egg reduction rate of repeated-dose versus single-dose mebendazole across two trials totaling 490 participants (246 repeated-dose, 244 single-dose; Steinmann et al., 2011^27^, weight 53.5%; Eshetu et al., 2020^28^, weight 46.5%). While both individual studies significantly favored repeated-dose treatment—Eshetu et al.^28^ with a mean difference of 0.31 (95% CI: 0.18 to 0.44) and Steinmann et al.^27^ with 0.09 (95% CI: 0.04 to 0.14)—the pooled random-effects estimate yielded a mean difference of 0.19 (95% CI: -0.02 to 0.41). Because this pooled interval spans zero, the overall difference between regimens is not statistically significant (Z = 1.75, p = 0.08). Significant statistical heterogeneity was observed (I² = 90%, Tau² = 0.02, Chi² = 10.16, df = 1, p = 0.001), indicating substantial inter-study variability that accounts for the wide pooled confidence interval. **(Figure 10)**

**Figure 10.**
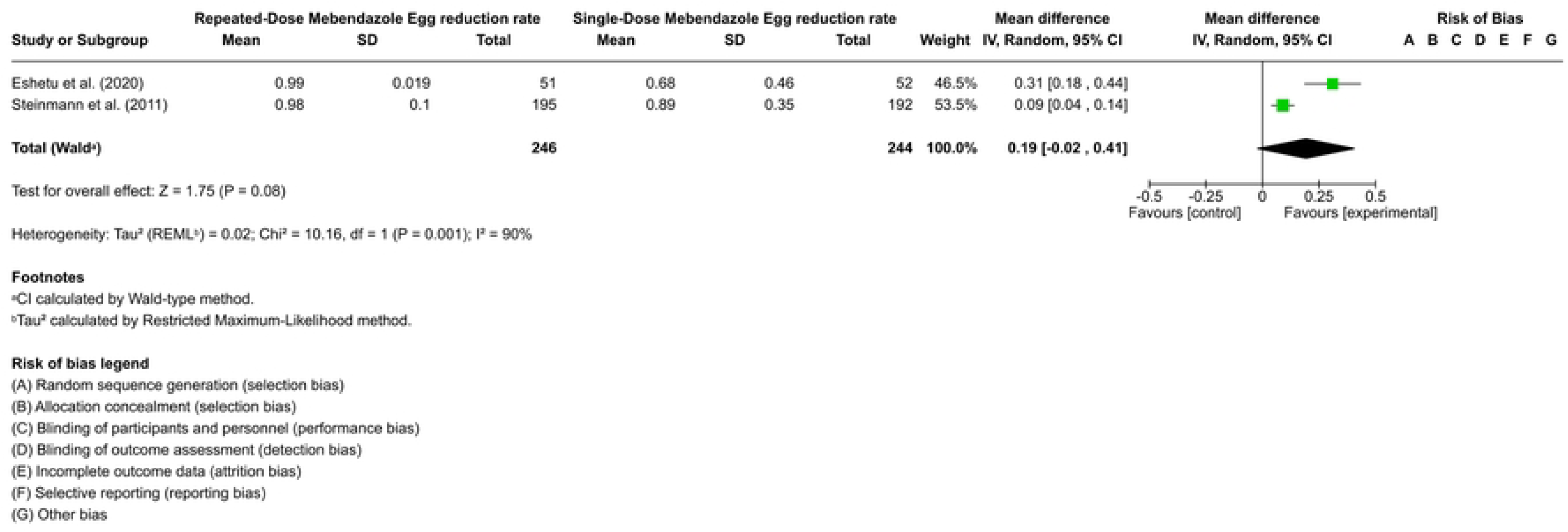
Forest plot comparing egg reduction rates of repeated-dose versus single-dose mebendazole

## Discussion

The comparative synthesis of our meta-analytic findings alongside the existing trial evidence highlights critical nuances regarding the efficacy of repeated-dose versus standard single-dose benzimidazole (Albendazole/Mebendazole) regimens in pediatric populations. Our overall pooled analysis demonstrated a marked superiority of repeated dosing in improving overall cure rates (OR 4.47, 95% CI: 1.58–12.67; post-sensitivity OR 2.60, 95% CI: 1.76–3.83) and egg reduction rates (MD 0.17, 95% CI: 0.01–0.32; post-sensitivity MD 0.08, 95% CI: 0.05–0.11).

When evaluating albendazole-based regimens, the evidence consistently supports the adoption of multi-day dosing to achieve optimal parasitological clearance. Our sub-analysis revealed that repeated-dose albendazole yields over three times higher odds of cure compared to single-dose therapy (OR 3.10, 95% CI: 2.17–4.44; I² = 7%), alongside a modest but statistically significant enhancement in egg reduction rate (MD 0.07, 95% CI: 0.03–0.11; I² = 0%). These findings closely mirror the clinical observations reported across individual trials. For instance, **Kim et al.**^25^ demonstrated in a factorial randomized trial among Ugandan schoolchildren that dual-dose albendazole significantly improved hookworm cure rates over single-dose administration (96.4% vs. 83.9%; OR 5.07, 95% CI: 1.61–15.96). Similarly, **Adegnika et al.**^24^ reported that while single-dose albendazole is sufficient for Ascaris lumbricoides (85% cure rate), extended dosing regimens are required to achieve adequate clearance against hookworm (92% cure rate after two doses) and Trichuris trichiura (83% cure rate after three doses). **Steinmann et al.**^27^ likewise underscored the necessity of a triple-dose albendazole regimen to attain acceptable therapeutic endpoints against A. lumbricoides and hookworm in high-endemicity settings.

In contrast, the comparative synthesis for mebendazole reveals substantial inconsistency and lack of definitive therapeutic superiority across multi-dose trials. Our meta-analysis yielded a wide, non-significant pooled odds ratio for mebendazole cure rates (OR 4.52, 95% CI: 0.44–46.24; I² = 95%) and a non-significant pooled egg reduction rate difference (MD 0.19, 95% CI: -0.02 to 0.41; I² = 90%). This marked heterogeneity stems from conflicting outcomes reported in individual clinical trials. While **Eshetu et al.**^28^ observed a dramatic therapeutic gain with multiple-dose mebendazole against hookworm infections in Ethiopia (OR 55.13, 95% CI: 11.92–255.00), other high-quality trials offer a much more conservative view. **Steinmann et al.**^27^ noted only a moderate benefit for repeated mebendazole dosing, whereas **Namwanje et al.**^26^ demonstrated that single and double doses of mebendazole provided essentially equivalent, low cure rates against T. trichiura in Ugandan schoolchildren (OR 0.95, 95% CI: 0.38–2.34). These conflicting observations indicate that mebendazole efficacy is highly sensitive to local species distribution, baseline infection intensity, and potential bioavailability limitations, making multi-dose mebendazole less reliable as a universal therapeutic upgrade compared to albendazole.

From a public health and policy perspective, these findings present a critical trade-off between logistical feasibility and clinical efficacy in preventative chemotherapy programs. World Health Organization guidelines currently advocate for single-dose mass drug administration (MDA) with albendazole or mebendazole due to low operational costs and high community compliance. As noted by **Adegnika et al.**^24^, single-dose campaigns remain attractive because they successfully reduce overall worm burdens and heavy multi-species parasite loads. Furthermore, **Kim et al.**^25^ pointed out that although co-administration with fatty foods (such as avocado) does not yield significant gains in hookworm clearance, expanding dosing frequency remains a primary, viable mechanism to enhance drug efficacy and delay the selection pressure for benzimidazole resistance. However, as demonstrated by the pooled data across all included trials, relying solely on single-dose preventive chemotherapy leaves a substantial reservoir of persistent infections, particularly for T. trichiura and hookworm.

## Conclusion

Repeated-dose albendazole demonstrates clear superiority over standard single-dose regimens for pediatric soil-transmitted helminth infections, significantly increasing cure and egg reduction rates. Conversely, multi-dose mebendazole shows variable, non-significant gains. Transitioning mass drug administration programs to targeted, multi-dose albendazole protocols—rather than mebendazole—offers an effective public health strategy to eliminate residual parasite reservoirs in high-transmission endemic regions.

## Recommendation

Based on our meta-analytic findings:

1. Shift from Universal Single-Dose to Targeted Multi-Dose Albendazole
2. Standardize Drug Selection Prioritizing Albendazole over Mebendazole
3. High-density or persistently endemic hotspots must be allocated resources for multi-day, school-based treatment campaigns.

## Data Availability

All relevant data are within the manuscript and its Supporting Information files. No new primary dataset was generated all extracted data were derived from previously published studies cited in the article.

